# Quantification of Covid-19 Vaccine Coercion in India: A Survey Study

**DOI:** 10.1101/2023.07.24.23293089

**Authors:** Bhaskaran Raman, Amitav Banerjee, Sai Mahesh Vajjala

## Abstract

**Introduction:** Informed consent is the cornerstone of medical ethics, enshrined in the constitution of most countries, as well as in international documents. However, mandates for Covid-19 vaccination as well as coercion was prevalent in many places in the world, including in India. Against this background, we did a cross sectional study to assess and quantify the extent of Covid-19 vaccine coercion in India.

**Methods:** A cross sectional study was conducted after obtaining ethical clearance from IIT Bombay. This survey was conducted using a pretested questionnaire anonymously amongst the college students and adults in Mumbai from October 2022 to December 2022. The questionnaire contained details of why the vaccine was taken, and if the participant was a student. Descriptive analysis was conducted and frequencies, percentages along with 95% confidence intervals were used to summarize the findings.

**Results:** A total of 483 participants responded, which included both students and non-students, of which 470 participants reported having taken the vaccine. A total of 106 (21.95%, 95% C.I. 18.48%-25.85%) reported to have pressured into taking the vaccine. The level of coercion was similar among college students 78 (21.61%, 95% C.I. 17.67%-26.14%) and non-student adults 28 (22.95%, 95% C.I. 15.82% – 31.43%).

**Conclusion:** A significant proportion was coerced into taking the vaccines, violating the requirement for informed consent. These results are of paramount importance for future policies as well as for posterity.

## Introduction

Informed consent is of paramount importance in medical ethics. The right to one’s health and bodily autonomy is guaranteed in Article-21 of the Indian constitution. Informed consent is also given unequivocal importance in the international UNESCO declaration of bioethics in the article 6. [1]. However, policies related to COVID-19 vaccine administration have been coercive in nature around the world, including in India. Covid-19 vaccine mandates for various aspects of day-to-day life were common in 2021 and early 2022. Few such examples in India are as follows, i) at workplace in Tamil Nadu state [2–4], ii) for accessing public transportation in Maharashtra state [5–7], iii) for accessing government services in Gujarat state [8–10], even the high court had upheld the decision of covid vaccine mandate by the Ahmedabad Municipal Commissioner in this case, iv) for entry into malls in Maharashtra state [6,7], v) for obtaining the ration from Public distribution System in the Madhya Pradesh state [11,12], vi) to enter educational institutions like colleges in Karnataka state [13,14] and vii) schools in the Chandigarh Union Territory [15].

On 02 May 2022, the Indian Supreme Court in the judgement of the case, “**Jacob Puliyel vs Union of India”** in paragraph 3 of the conclusion, ruled that such coercion is disproportionate and violative of the Indian constitution, especially Article 21. [16,17] Further they opined that bodily integrity is protected under Article 21 of the Constitution and no individual can be forced to be vaccinated. [16,17] While the Hon’ble Supreme Court ruled on the unconstitutional nature of the mandates, various forms of mandates and coercion continued for some time.

We did a cross sectional study to ascertain the extent of this coercion. This study will be an important input in future policies, and also for historical record.

## Survey Methodology

The survey sought to quantify the extent of Covid-19 vaccine coercion among students as well as non-student adults. Since the survey involved personal health information, it was designed to be anonymous: name or other identifying information was not collected or even asked during the survey. In the student category, only college students were considered, not school students. College students and adults who were willing to participate in the study were chosen as study participants. In order to obtain this sensitive information and to maintain confidentiality, the data was collected anonymously. After obtaining the Institutional Ethics Board approval, data collection was started.

15 sessions were conducted on different dates between 26^th^ October 2022 and 19^th^ December 2022 at two prominent sets of locations: within college campuses and outside. While the former primarily targeted the student category, the latter primarily targeted the non-student category. Within college campuses, various venues such as food court, hostel, classroom were considered. Outside of college campuses, various locations like bus-stops, markets, and local train stations were considered. All survey locations were in the city of Mumbai, Maharashtra, India.

The following statement of informed consent was shown to a potential survey participant.

### Purpose of study

*Informed consent is of paramount importance. Several Covid-19 vaccine policies have been coercive, and there is even a Supreme Court ruling (02 May 2022) that such coercion is disproportionate and violative of the Indian constitution. This study seeks to measure the effect of the coercive policies, as it will be an important input in future policies, and also for historical record*.

#### In this survey, you are asked 2 questions

*No personal information is collected or stored. The survey results will be summarized for students and non-students separately. It may be published, to improve understanding of past policies. Please provide your answer to the above question after reading and understanding this consent and question form*.

The informed consent statement was shown in English as well as in Hindi and explained briefly to a potential participant. The response was collected from those who agreed to participate in the survey. The two questions asked. For simplicity, there was exactly one question of content asked in the survey: *Q1 What is the PRIMARY reason you took the Covid-19 vaccine?* The participant had to choose from among six choices: *A) Willingly, for own health; B) Willingly, for others’ health; C) Pressured to take, for travel; D) Pressured to take, for education or work; E) Pressured, for some other reason; F) Not taken the vaccine*. The first two responses would be considered as willingly vaccinated while the latter three would be considered as vaccine coerced for analysis. Since we wanted to separate the survey results by students versus non-students, an additional logistical question asked was: *Q2 Are you a college student? Yes or no as answer*.

### Sample size estimation

To estimate an assumed proportion of 20% vaccine coercion among the college students and adults, at an acceptable error of 4% with alpha error at 5% and 80% power the minimum sample size required would be 385. The software used was WinPepi v11.65. However, we could collect a higher number of responses: 483 responses, of which 470 participants reported having taken the vaccine.

Data collected using both the techniques were entered into Microsoft Excel sheet and analyzed using Epi Info software developed by the Centers for Disease Control and Prevention. Descriptive statistics were applied. Frequency, percentages and 95% Confidence Intervals were used to summarise the collected data.

### Institute Ethics Committee (IEC) Approval

This survey study was approved by the Institute Ethics Committee (IEC) of the Indian Institute of Technology Bombay (IITB), on 18 October 2022. Approval number: IITB-IEC/2022/026.

## Results

A total of 483 valid responses could be recorded. This consisted of 361 students and 122 non-student adults. Among which 13 participants did not take the vaccine. Further analysis showed that, for non-student adults, 28 (22.95%, 95% C.I. 15.82% -31.43%) [Table 1] reported that the reason for taking the vaccine was pressure due to travel, education or others which was similar in college students as well where 78 (21.61%, 95% C.I. 17.67% - 26.14%) [Table 2] reported the same. Overall, 106 (21.95%, 95% C.I. 18.48% - 25.85%) [Table 3] reported that pressure as a reason for taking the vaccine. Figure 1 and Figure 2 summarize the survey results for non-students and students respectively. This clearly shows that a significant proportion of participants reported pressure as a reason for taking the vaccine. A small proportion of adults did not take vaccine in non-college adults, 5 (4.1%) which was lower than that among college students 8 (2.2%).

**Table 1:**
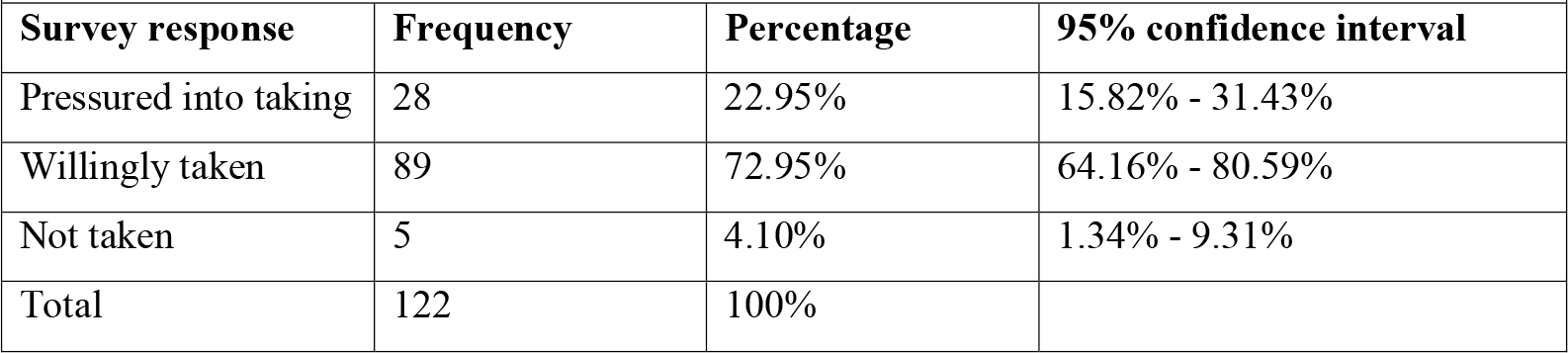
Responses from non-student adults.

**Table 2:** Responses from college students.

| Table 2: Responses from college students |  |  |  |
| --- | --- | --- | --- |
| Survey response | Frequency | Percentage | 95% confidence interval |
| Pressured into taking | 78 | 21.61% | 17.67% - 26.14% |
| Willingly taken | 275 | 76.18% | 71.52% - 80.28% |
| Not taken | 8 | 2.22% | 1.13% - 4.31% |
| Total | 361 | 100% |  |

**Table 3:** Responses from total participants (students & non-students)

| Table 3: Responses from total participants (students & non-students) |  |  |  |
| --- | --- | --- | --- |
| Survey response | Frequency | Percentage | 95% confidence interval |
| Pressured into taking | 106 | 21.95% | 18.48% - 25.85% |
| Willingly taken | 364 | 75.36% | 71.33% - 79% |
| Not taken | 13 | 2.69% | 1.58% - 4.55% |
| Total | 483 | 100% |  |

**Figure 1:**
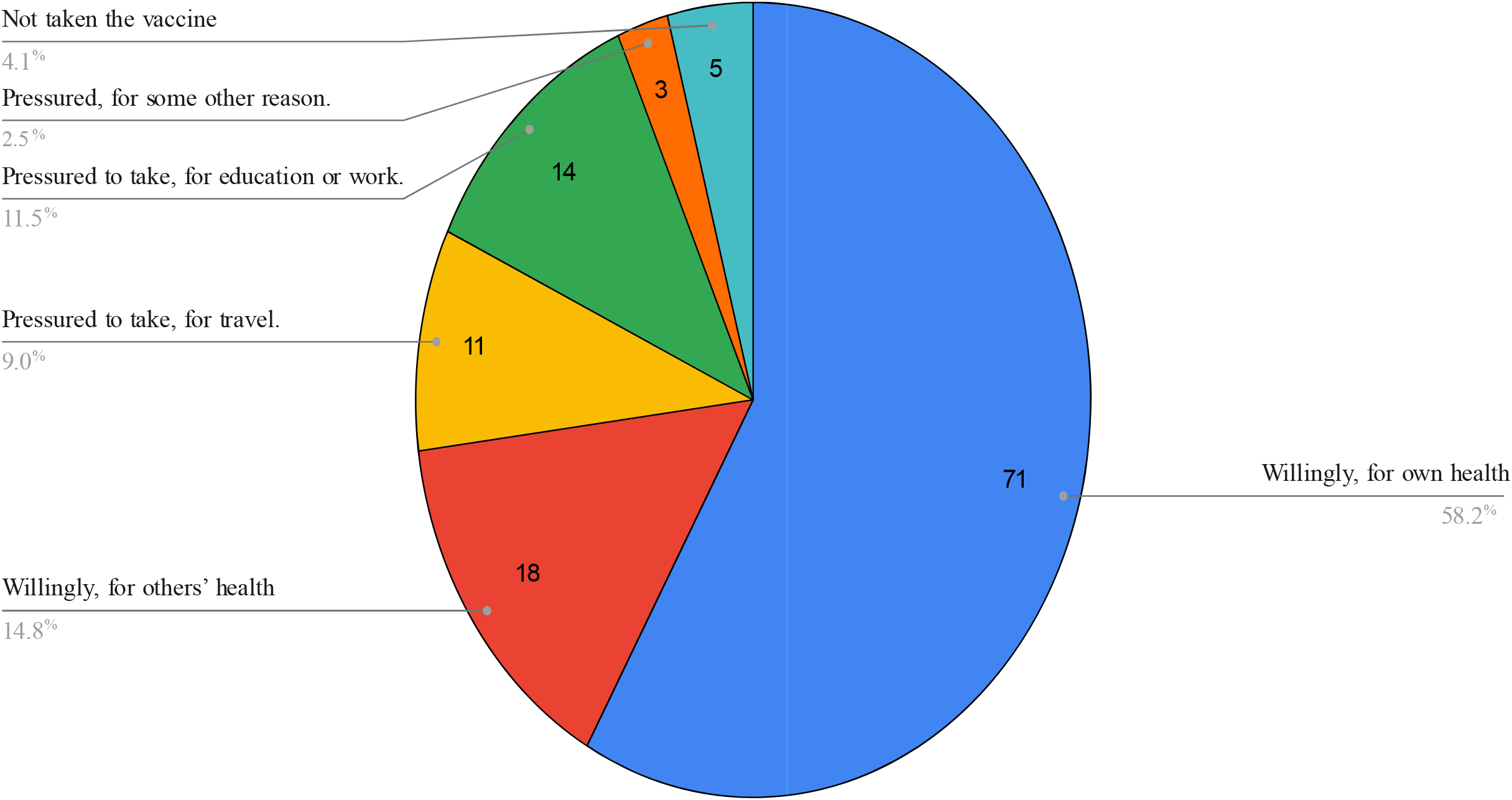
Pie chart of responses from non-students (total: 122)

**Figure 2:**
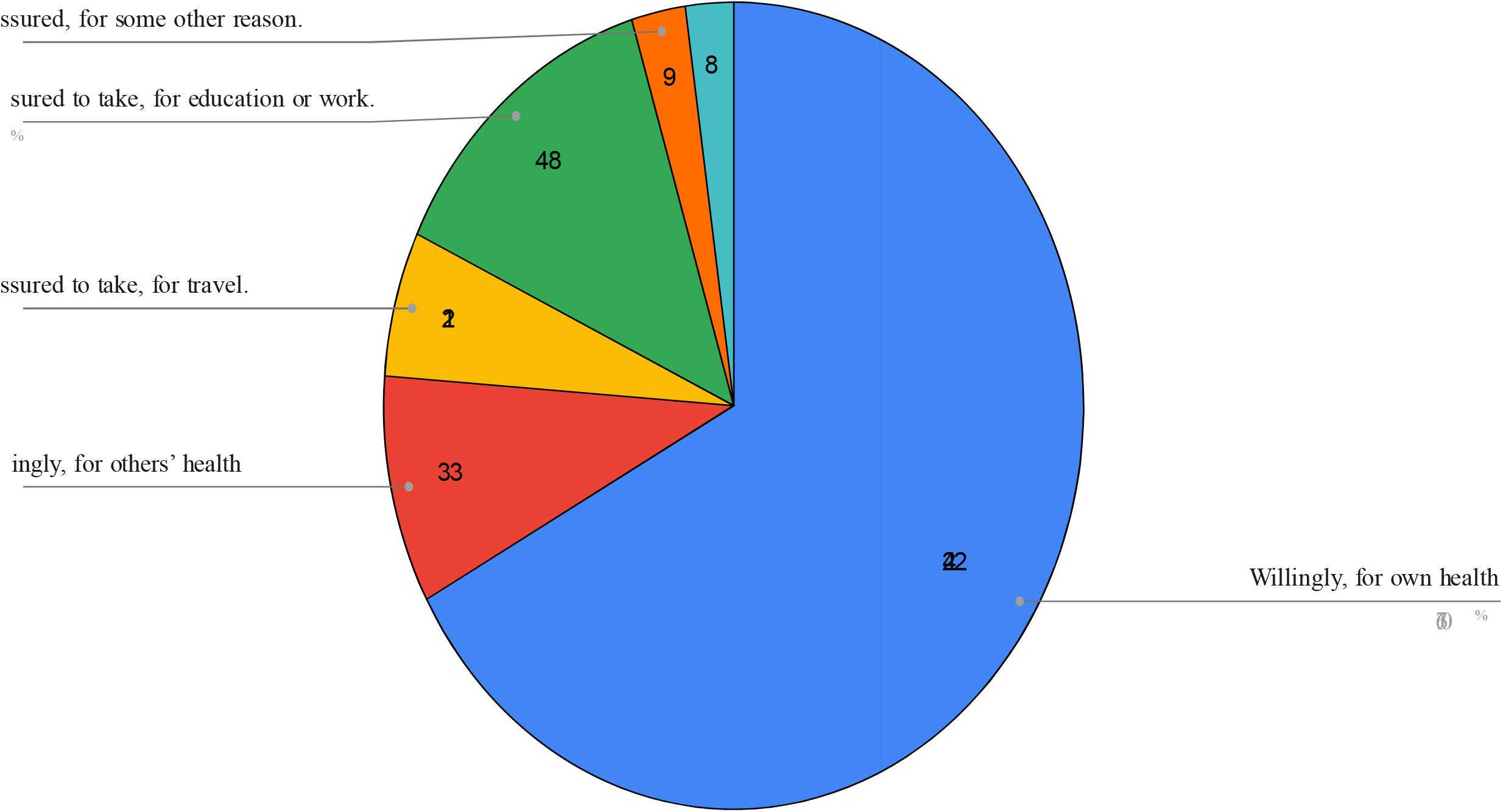
Pie chart of responses from student (total: 3)

## Discussion and Conclusion

A significant percentage of students as well as non-students, about 1 in 5, reported that they have been coerced into taking the Covid-19 vaccine. This is significant, especially given the Supreme Court ruling of the unconstitutionality of the vaccine mandates. The Government of India has claimed in its affidavit in another Supreme Court case that Covid-19 vaccination is voluntary [18]. Although the vaccination was voluntary as per the union government, our survey finds that due to various coercive policies on the ground, a large section of the population was pressured into taking the Covid-19 vaccines. This has important implications for future policies, as it concerns the issue of trust in public health.

The coercive policies for the college student population are especially significant, as data from the US as well as Europe shows that this age-group had no excess deaths in 2020 or 2021, due to Covid-19, a mortality rate of 0.03 per 1,00,000 in age group 0-19 years [19]. Even during the peak of August 2021 to September 2022, study from US revealed that COVID mortality was 0.6 per 100 000 for those aged 1 to 4 years; 0.4 per 100 000 for those aged 5 to 9 years; 0.5 per 100 000 for those aged 10 to 14 years; and 1.8 per 100 000 for those aged 15 to 19 years. [20]

The survey study has been limited in scope, and it is difficult to extrapolate the results to other places such as rural regions. In other places, the level of coercion could have been lower or higher. The background details of the study participants were not taken which would provide more accuracy in the estimation. Qualitative studies on coercion can elaborate the understanding on vaccine coercion and would generate insights into the policy makers as well as healthcare givers. This observational descriptive study provides an insight into vaccine coercion and brings this into light which requires further studies to fill the lacunae and establish stronger evidence.

## Data Availability

All data produced in the present study will be made available upon reasonable request to the authors

## Supplementary file

